# DESIGNING, DEVELOPING AND VALIDATING A SET OF STANDARDIZED PEDIATRIC PICTOGRAMS TO SUPPORT PEDIATRIC-REPORTED GASTRODUODENAL SYMPTOMS

**DOI:** 10.1101/2023.08.14.23294049

**Authors:** Gayl Humphrey, Celia Keane, Armen Gharibans, Christopher N. Andrews, Alain Benitez, Hayat Mousa, Gregory O’Grady

## Abstract

**Objective:** To develop and validate a set of static and animated pediatric gastroduodenal symptom pictograms.

**Methods:** There were three study phases: 1: Co-creation used experience design methods resulting in ten pediatric gastroduodenal symptom pictograms (static and animation); 2: an online survey to assess acceptability, face and content validity; and 3: a preference study. Phases 2 and 3 compared the novel paediatric pictograms with existing pictograms used with adult patients.

**Results:** Eight children aged 6-15 years (5 Female) participated in Phase 1, 69 children in Phase 2 (median age 13 years: IQR 9-15), and an additional 49 participants were included in Phase 3 (median age 15: IQR 12-17). Face and content validity were higher for the pediatric and animated pictogram sets compared to pre-existing adult pictograms (78% vs. 78% vs. 61%). Participants with worse gastric symptoms (lower PedsQL-GIS score) had superior comprehension of the pediatric pictograms (χ^2^_8_ < .001). The pediatric pictogram set was preferred by all participants over animation and adult (χ^2^_2_ < .001).

**Conclusion:** The co-creation phase resulted in the symptom concept confirmation and design of ten acceptable static and animated gastroduodenal pictograms with high face and content validity when evaluated with children aged 6 to 18. Validity was superior when children reported more problematic symptoms. Therefore, these pictograms could be used in clinical and research practice to enable standardized symptom reporting for children with gastroduodenal disorders.

**Why is it important:**

▪ Diagnosis of gastroduodenal disorders of the gut-brain interaction (DGBI) in pediatrics is difficult as symptoms often overlap.
▪ Pediatric patients find identifying and distinguishing symptoms difficult.
▪ Validated gastroduodenal symptom pictograms have been found to help adults accurately report their symptoms and have been used effectively to standardize symptom monitoring, including continuous symptom reporting during investigations.
▪ There are no validated pediatric gastroduodenal symptom pictograms.

**What we did:**

▪ Co-created a set of ten pediatric gastroduodenal symptom pictograms.
▪ Undertook a face and content validity study to assess the novel pictograms with 118 pediatric participants with a median PedsQL-GIS score of 86.1 (IQR 68.1-90.0).

**The Outcome:**

▪ Designed a novel set of pictograms with face and content validity that were preferred over other sets, enabling acceptable, simple and validated pediatric patient reporting of their gastroduodenal symptoms.

## INTRODUCTION

Gastroduodenal disorders of the gut brain interaction (DGBIs) are a group of disorders where patients experience an impairment in gastroduodenal function that a biochemical, structural or organic abnormality cannot explain. These disorders result in high healthcare usage in children and young people (1). Clinical evaluation is challenging as a diagnosis is based on symptom criteria, but individual disorders do not have discrete symptomatology. Abdominal pain, nausea, vomiting, epigastric bloating, reflux and postprandial fullness are commonly reported symptoms of gastroduodenal DGBI.(2, 3) Patient reporting of symptoms can be further complicated in children who may find identifying and differentiating their symptoms difficult or find the terms used to describe symptoms, such as early satiety, challenging to comprehend.

Validated pediatric assessment instruments such as the PedsQL™ Gastrointestinal Symptom Scales (3) and the Rome IV Criteria for the Diagnosis of Functional Gastrointestinal Disorders in Children (2) aid clinical diagnosis. A parent-proxy instrument is often used in place of self-completion, as a high degree of literacy and cognitive capability is required to navigate the questionnaires. This can introduce a degree of perceptual inaccuracy, and some report low concordance in symptom severity between a child and their proxy’s reporting. (4, 5) Therefore, a simpler patient-reported instrument is required to enable pediatric patients to report their symptoms more accurately. Ideally, this tool could be used statically and dynamically (for continuous reporting) in association with diagnostic testing and for repeated assessments to evaluate the efficacy of interventions.

Visual representations of gastrointestinal symptoms as pictograms have been used to overcome some of the difficulties of longer assessment instruments and can enable more immediate reporting. (6) They have been found to improve understanding when a concept is challenging to express in words or when the terms used may be a barrier to comprehension. Children become familiar with visuals from an early age as a way to help them with comprehension (6), making pictograms an ideal medium for a diverse range of children and young people. A set of gastroduodenal symptom pictograms has been developed for adults with DGBI. (7) These pictograms were subsequently shown to have high convergent validity and high concurrent validity with the patient assessment of the upper gastrointestinal symptom severity index (PAGI-SYM) (8, 9) and have been used effectively for dynamic symptom reporting alongside gastric electrophysiology testing. (10) The successful use of pictograms for symptom reporting in adults presents an opportunity to develop a similar system for use in pediatric populations.

This study aimed to design and validate a set of pictograms to support paediatric patients reporting gastroduodenal symptoms. There were three study phases (Figure 1).

**Figure 1.**
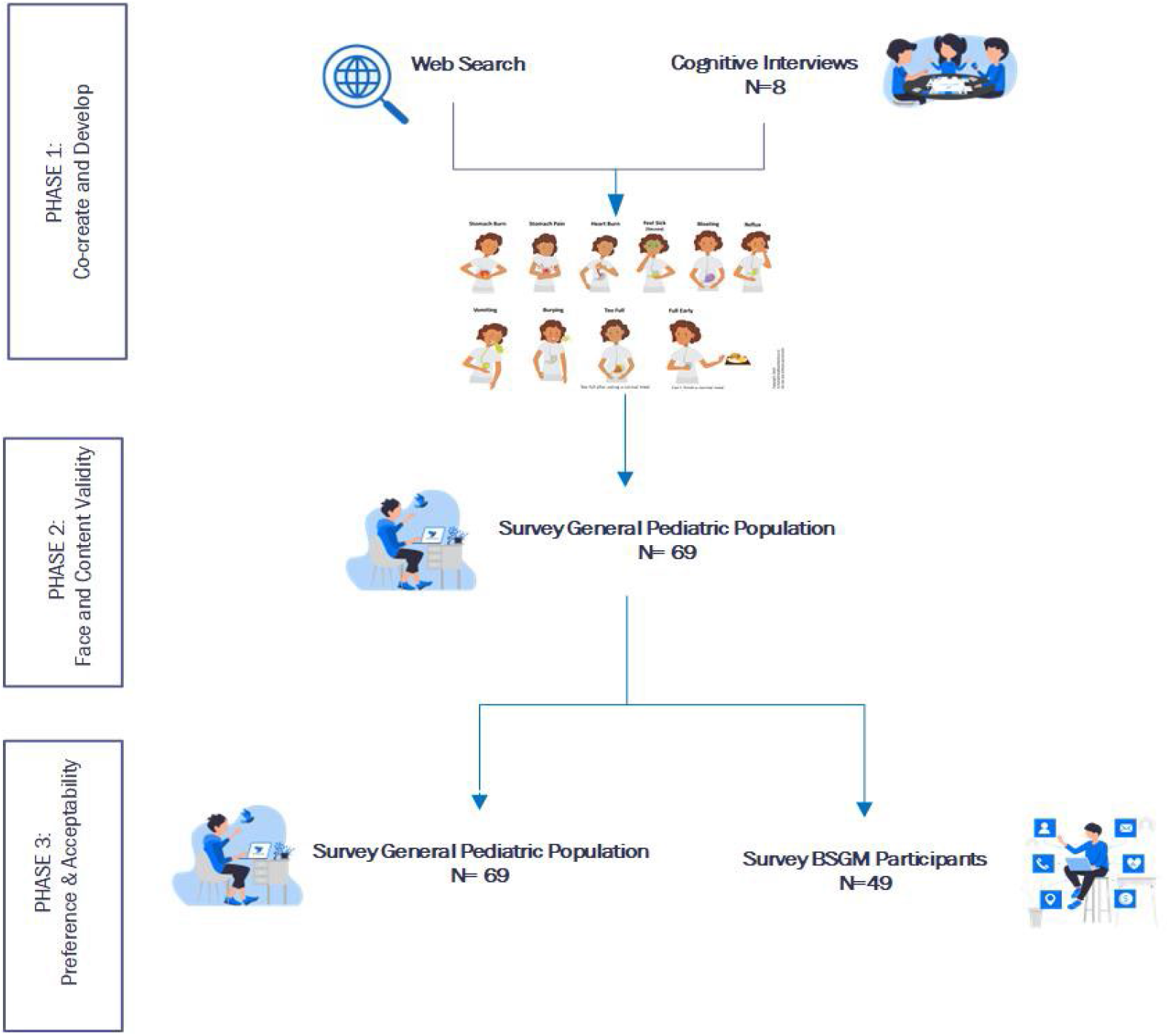
Pictogram Study Phases

Specific study objectives were to,

1. Co-design a set of comprehensible, acceptable, and relatable gastroduodenal symptom pictograms with and for children and young people.
2. Assess face and content validity of the pictograms, including,
  a. Comparison between the Pediatric pictograms with validated Adult pictograms (Figure 2);
  b. Explore differences in face and content validity between 5-12 years and 13-18 years age groups;
  c. Explore the relationship between the presence of symptoms and face/content validity.
3. Assess pictogram set preference between the validated adult and new pediatric animation and static pictograms.

**Figure 2:**
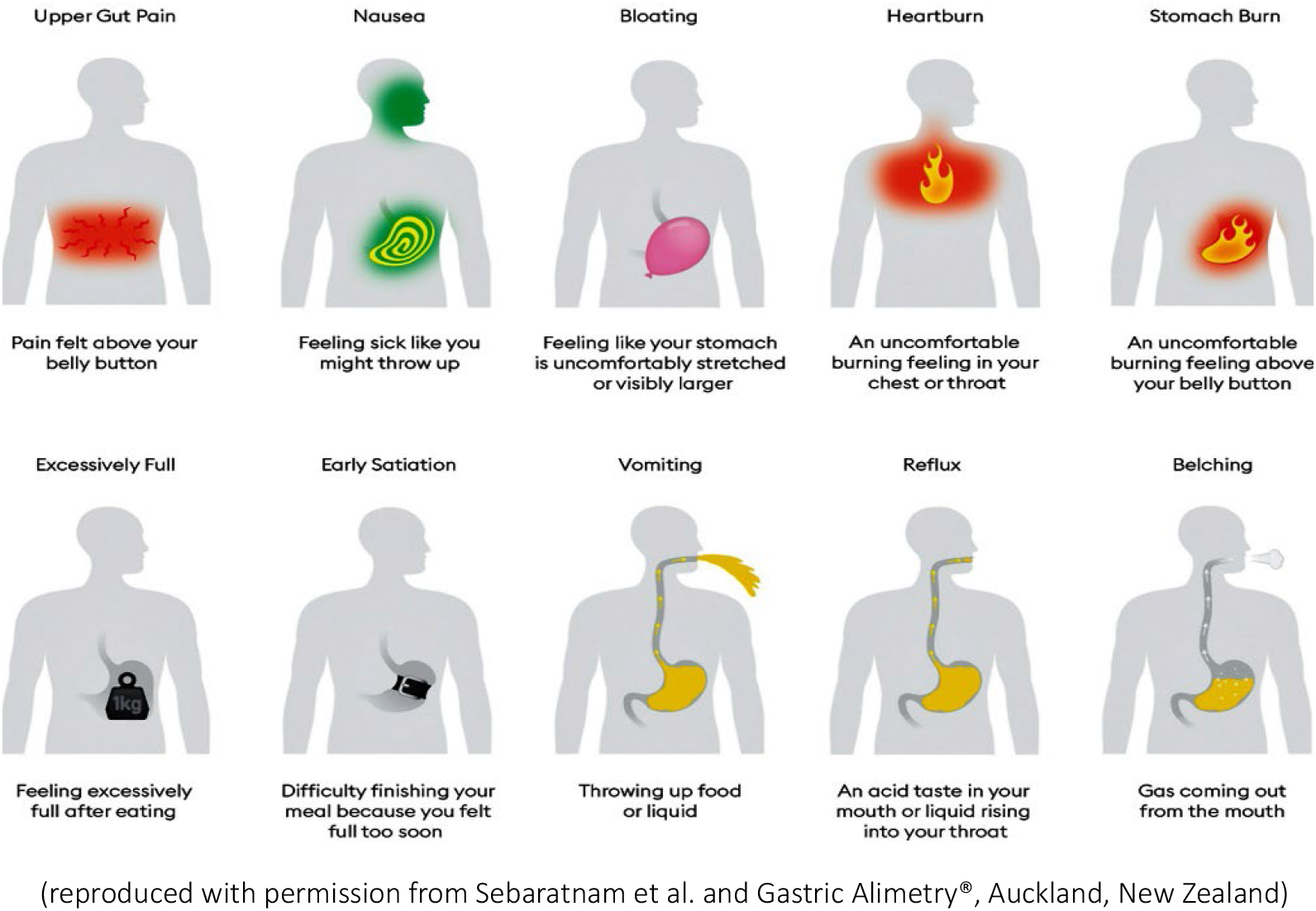
Validated Adult Pictograms

## MATERIALS AND METHODOLOGY

The New Zealand Health and Disability Ethics Committee granted ethical approval (2022 AM 12705). Participation was voluntary, and informed consent or assent, with parental consent, was obtained.

This study uses a mixed methods approach. It adopted the International Organisation for Standardisation (ISO 9186-1 (11) and ISO 9186-2 (12)) methodology for face validity and pictogram preference studies. The digital tools used to collect data are Health Insurance Portability and Accountability Act (HIPAA) compliant, and reporting follows the CHERRIES statement for web-based questionnaires. (13)

Participants completed demographic questions and the Pediatric Quality of Life-Gastrointestinal Symptom questionnaire (PedsQL-GIS) (14) to measure health-related quality of life related to the occurrence and impact of gastrointestinal symptoms.

### Phase 1. Co-creation of Pediatric Pictograms

#### Web-Search

Two web image searches were undertaken using search terms underpinned by the cardinal symptoms of gastroparesis and the Rome IV Functional Dyspepsia symptom criteria (2, 15) to identify images available in the public domain. Image atributites were collected, including symptoms portrayed, colors, facial expressions, text/descriptions, body language, props, and abstract lines to indicate context, emotion, or movement; face, part of, or whole body displayed.

#### Pediatric Interviews

Participants with experience of gastric symptoms such as nausea, abdominal pain and vomiting but not necessarily a gastroduodenal DGBI diagnosis were identified through networks and invited to participate in two rounds of interviews. Two rounds of qualitative interviews were then undertaken with the children aged 5-18 years, including cognitive probing, to evaluate the understanding of the symptom names and descriptions. During these interviews, the validated adult pictogram names and descriptions (7, 16) were used: bloating, belching, excessively full, early satiety, heartburn, nausea, stomach burn, vomiting, and upper gut pain. These were selected as they constituted the typical gastroduodenal symptoms identified via consensus methodology and the accumulation of epidemiological and pathophysiological data. (2, 15, 17)

In Round 1, the symptoms were described to participants, the terms used were clarified, and alternatives were captured. Once participants were comfortable with the symptoms, they were asked to draw a picture of each symptom and give a running commentary about what they drew and what made the image and its elements meaningful. Based on the web search and the initial interview findings, one author developed a draft set of ten pediatric static and animation gastroduodenal pictograms.

In the second round of qualitative interviews, the draft Pediatric pictograms were presented to ascertain whether each pictogram was recognizable, understandable, acceptable and relatable. The symptom labels and descriptions were presented, and each participant was asked to select which term for each pictogram they found readily understandable.

Contemporaneous written notes were taken during each interview, and interviews were audio-recorded if participants consented. Drawings were either collected or photographed.

### Phase 2: Face and content validity: Comparison of the Pediatric (Static and Animation) pictograms to validated Adult pictograms

A convenience sample of the general pediatric population (reported as General) was recruited using flyers, social media, colleagues, and networks during 2022. Eligibility and inclusion criteria were that participants were aged 5-18 and living in Aotearoa | New Zealand.

An online survey was developed and tested using Qualtrics (Qualtrics, Provo, UT) (18) and accessed through a QR code. Eligible participants were directed to an animated video which presented the study information. The video was used to maximize appeal and understanding for younger participants and those with literacy challenges or where English may not be a first language. Aer consenting, demographic data and the PedsQL-GIS questionnaire were collected. The Internet Protocol (IP) address was captured for each completed questionnaire to check and remove multiple responses by a single user. Survey completion me was collected, and any completed within 10 mins were excluded.

Two rounds were used to determine content and face validity. Round 1 adopted a word-image-matching approach. A single symptom pictogram was presented with a set of text labels, including a ‘No idea’ response. Participants were prompted to choose the text label they thought best named the symptom shown by the pictogram. If a selection was not made within 30 seconds, it was assumed that the pictogram was not readily comprehended, and the digital platform presented the following pictogram. All pictograms and response options were presented in random order.

In Round 2, incorrect pictograms (incorrectly labelled or no idea or response) were represented with a label and brief text description. Participants were asked if the label and description improved their comprehension of the symptom. A text box was available for participants to comment on improving the pictogram, label, and text description.

### Phase 3: Pictogram Perceptual Quality and Preference

Participants from Phase 2 and eligible pediatric participants with and without a gastroduodenal DGBI from an associated study investigating Body Surface Gastric Mapping (BSGM) were enrolled in Phase 3. (19, 20) All participants completed a Pictogram Perceptual Quality and Preference survey to assess preference for the novel paediatric pictograms presented as static images (referred to as Pediatric), the animated paediatric pictograms (referred to as Animated) or the Adult pictograms (referred to as Adult). Each symptom pictogram (Pediatric, Animated, Adult) was randomly presented side by side, and each participant was asked to select the pictogram they preferred. Then, each pictogram set (all 10 symptoms) was presented, and participants were asked to select their preferred set: Pediatric, Animated or Adult.

### Data Analysis

An inductive thematic analysis (21) was adopted for Phase 1 to uncover key insights, unique perspectives and identify what symptoms were challenging to comprehend, describe and draw. Quantitative data from Phases 2 and 3 were analyzed in SPSS (v.28 Chicago, IL). Descriptive statistics were used to present the two participant cohort characteristics, including PedQL-GIS scores. Phase 2 and 3 cohorts are combined, and overall percentages illustrate the differences in age and age groups in comprehension and preference. Age Groupings were selected to correspond with broad developmental stages (22) and to reflect school groupings in New Zealand: 5-12 years primary/intermediate school and 13-18 years high school.

Pictogram comprehension outcomes were transformed into a dichotomous output (0=incorrect and no idea and 1 =correct), and pictogram preference categories were coded as 0 (not preferred) and 1 (preferred). The Chi-Square test of independence was used to test comprehension of pictograms and pictogram preferences. Pictogram comprehension analyses were i) age and comprehension of individual pictograms, ii) age and comprehension of a pictogram set (Pediatric, Animation, Adult), iii) Age Groups and pictogram set comprehension, and iv) PedsQL-GIS outcome and pictogram comprehension. Pictogram set preference analyses were iv) age and pictogram preference, v) age and pictogram set, vi) Age Groups and pictogram set and vii) PedsQL-GIS and pictogram set.

## RESULTS

### Phase 1. Co-Design

#### Web Search

The web-based image search was undertaken on 10^th^ September 2021. The first three results pages included pictures of people, anatomical images, cartoons and pictogram-style images representing the ten symptoms. A mix of colors, gestures, and action lines were commonly used to illustrate the symptom location, an emotion or movement (Figures 3 and 4). Props were occasionally used in association with cartoon images.

**Figure 3.**
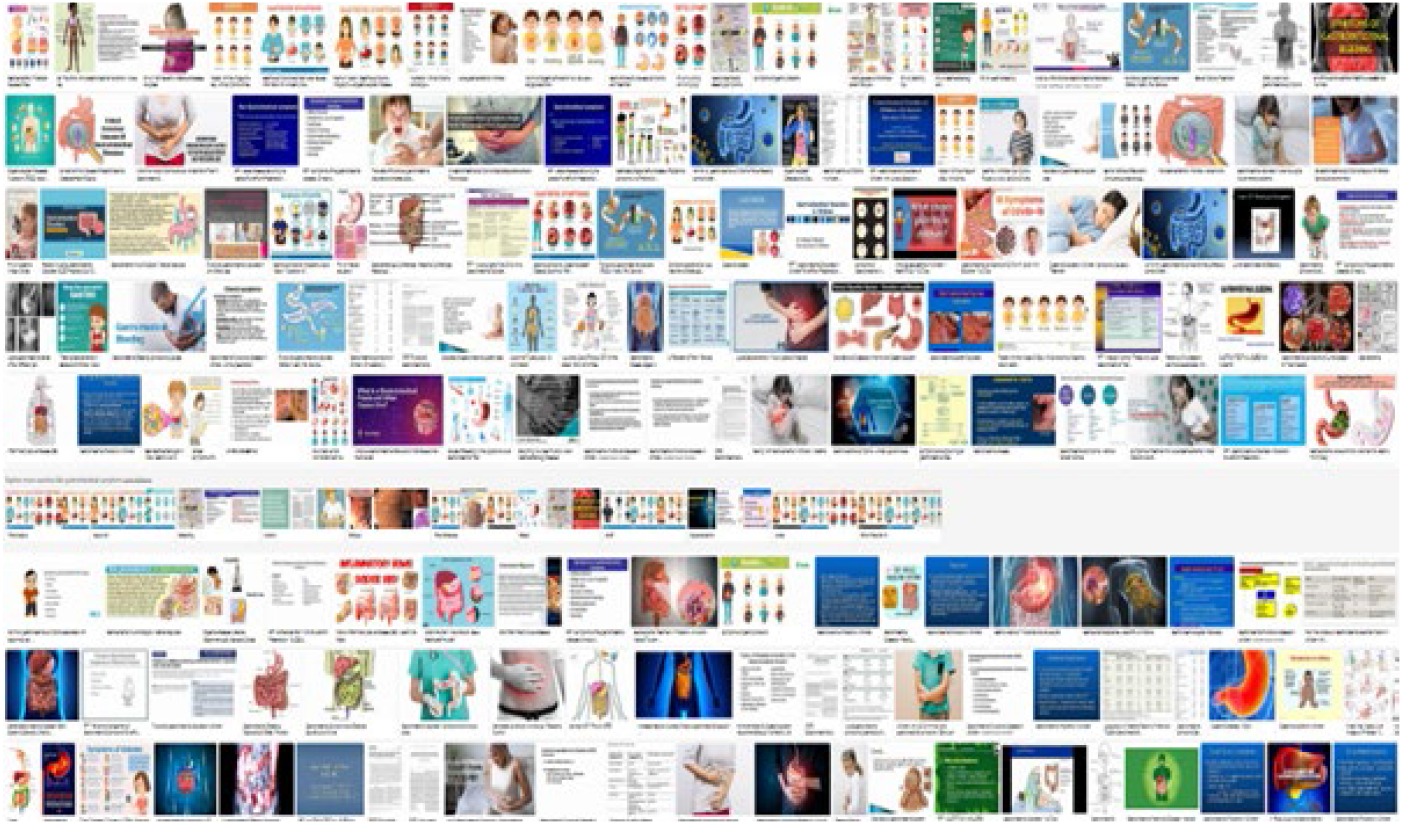
Example of a web search page outcome

**Figure 4.**
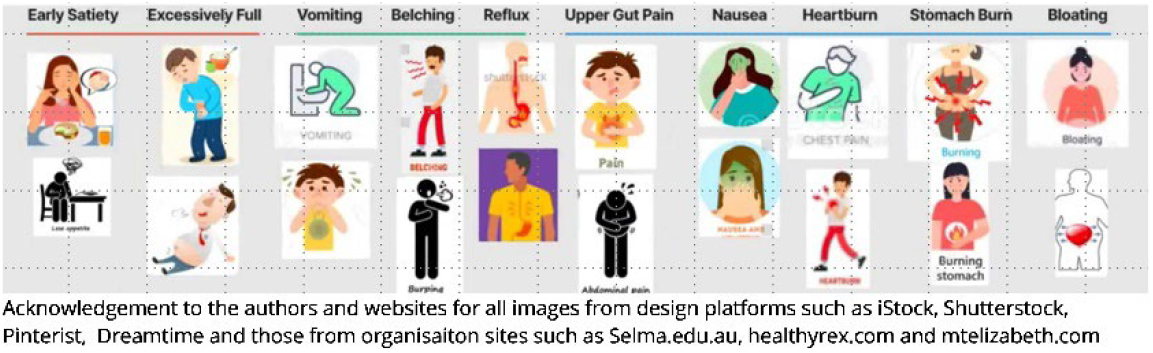
Pictogram style examples from web search by symptom type

#### Interviews

Eight children participated in the interviews: 3 males (ages 5-15) and 5 females (ages 9-15). Participants were NZ European (n=5), Fijian Indian (n=1), European (n=1), and Australian (n=1). Participants used similar terms to describe each symptom. ‘‘*Feeling dizzy and blugh*”, “*feeling yucky*”, and “*feeling green*” were used to describe nausea; “*fiery tummy*”, “*hot burny pain in my stomach*”, and “*acidy burn*” were used to describe stomach burn and “*puking*”, “*being sick*” and “*throwing up*” as an alternative to vomiting.

Participant drawings included a mix of emoji-style, partial body and full-body images (Figure 5). Participants added props to help recognize complex symptoms; for example, three participants had a plate of food barely touched next to the person to convey early satiety. Two participants had the person lying down “*because your tummy is so full, you can’t move*” to convey excessively full. Green was commonly used to illustrate nausea and vomiting, and red for upper gut pain. Four participants drew lightning-style lines to illustrate the intensity of the upper gut pain.

**Figure 5.**
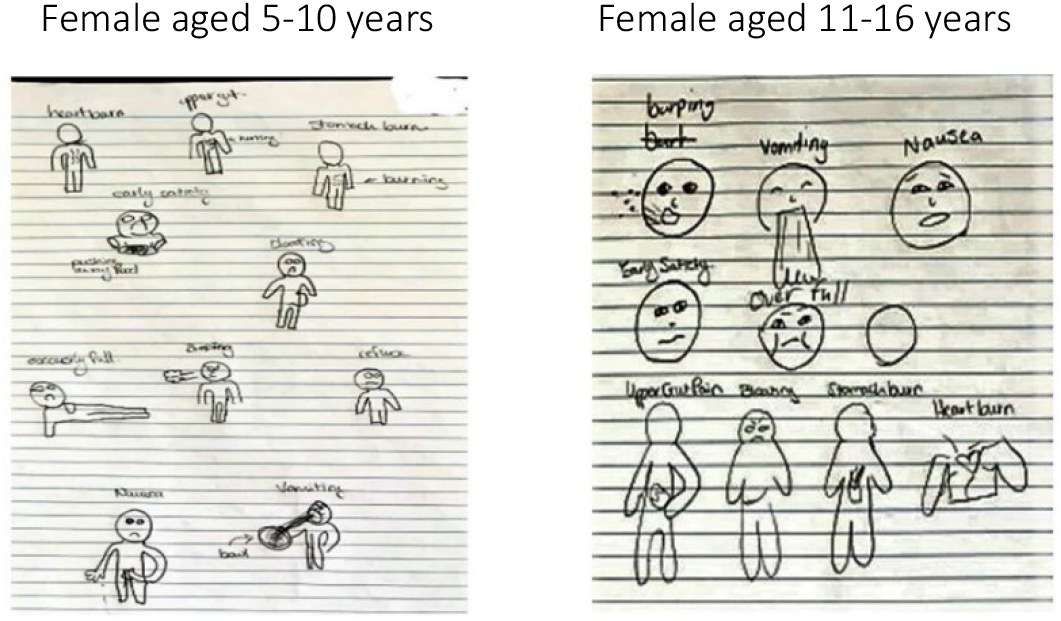
Examples of pictures of various gastroduodenal symptoms drawn by two female participants (aged between 5-10, and 11-16); the bowl with the vomiting image, a balloon prop for bloating, lines or a cloud shape near the mouth for belching, and the heart shape with wriggly lines around it for heartburn.

A draft set of static (Figure 6) and animated pediatric symptom pictograms were developed using the participant interview data and the web search findings.

**Figure 6.**
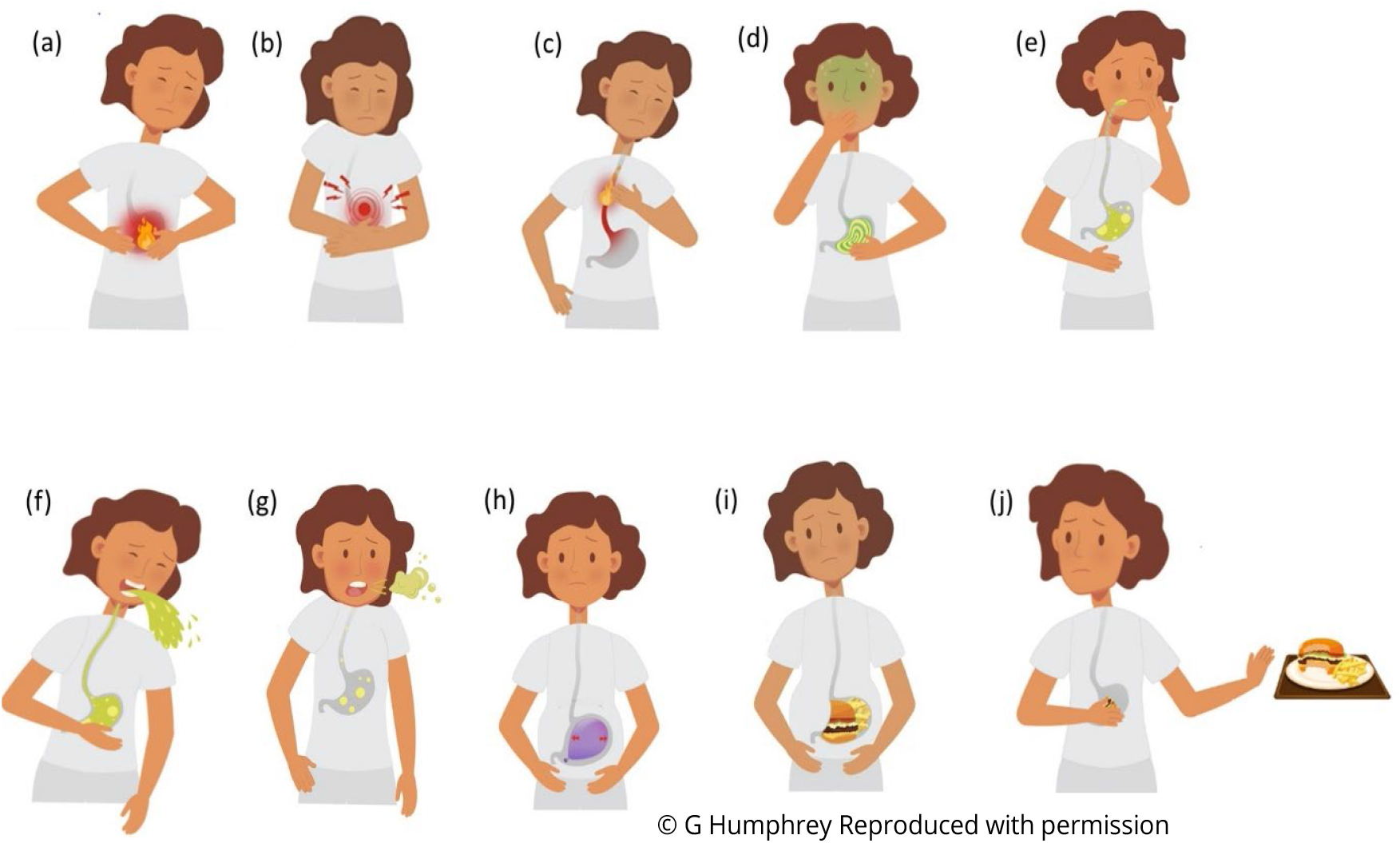
Pediatric gastroduodenal symptom pictograms. (a) stomach burn, (b) upper gut pain, (c) heartburn, (d) nausea, (e) reflux, (f) vomiting, (g) belching, (h) bloating, (i) excessively full, and (j) early Satiety. (The same images were used for the Animation set).

In Round 2 of qualitative interviews, participants responded positively to the draft pictograms and remarked that the images reflected what they drew and discussed during the first interview. The use of props, such as food in the stomach or the plate with the hand up, were;

> “…*like we discussed. They look great, and the colors, faces and that, make it easy to get what the problem, you know, umm, what each symptom is…”* (male, 11-16 years).

Another participant was excited to see the ideas she had contributed depicted in the pictograms and remarked that;

> *“…all of them are easy to understand. Everyone should be able to get these*.*”* (female, 5-10 years)

The depiction of the person in the pictogram was well-liked and reported as very relatable by all participants. There were no suggestions on specific areas for improvement, so there were no further rounds of iteration. However, the younger participants (≤ 10 years) suggested animating the pictograms;

> “…*would make it even easier for me to get what the problem is, as some of those problems I am still not sure about*…” (male 5-10 years).

Regarding the symptom labels, simplification and using language that they (children and young people) would understand were the overarching themes that emerged (Table 1).

**Table 1.**
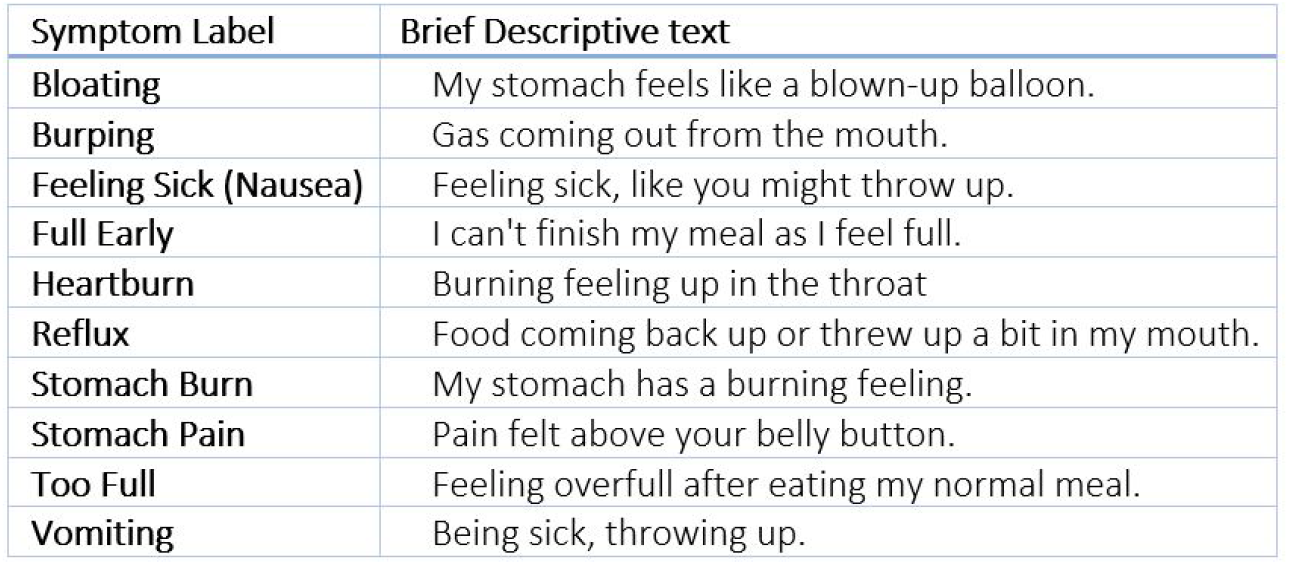
Suggested symptom labels and descriptors.

| Symptom Label | Brief Descriptive text |
| --- | --- |
| Bloating | My stomach feels like a blown-up balloon. |
| Burping | Gas coming out from the mouth. |
| Feeling Sick (Nausea) | Feeling sick, like you might throw up. |
| Full Early | I can't finish my meal as I feel full. |
| Heartburn | Burning feeling up in the throat |
| Reflux | Food coming back up or threw up a bit in my mouth. |
| Stomach Burn | My stomach has a burning feeling. |
| Stomach Pain | Pain felt above your belly button. |
| Too Full | Feeling overfull after eating my normal meal. |
| Vomiting | Being sick, throwing up. |

### Phase 2: Pictogram Face and Content Validity

Sixty-nine participants in Phase 2 visited the electronic survey site, and all completed the questionnaire with none excluded for missed data or completion quicker than 10 minutes: see Table 2 for demographics and PedsQL-GIS total and symptom domain scores. Relatively high median total PedsQL-GIS scores are not unsurprising, given that having gastroduodenal symptoms or a diagnosis of a DGBI was not an eligibility criterion for participation.

**Table 2.** Participant Demographics and PedsQL-GIS Scores.

|  |  | General Pediatric<br>n= 69 | BSGM<br>n= 49 | *p* | Combined<br>N=118 |
| --- | --- | --- | --- | --- | --- |
| **Sex**<br>**n (%)** | Female | 24 (35) | 22 (45) | ns | 46 (38) |
|  | Male | 34 (49) | 27 (55) |  | 61 (52) |
|  | Non-binary | 4 (6) |  |  | 4 (3) |
|  | Prefer not to say | 7 (10) |  |  | 7 (6) |
| **Age** | Age, median (IQR) | 13 (9-15) | 15 (12-17) | ns | 13.5 (9-16) |
| **Ethnicity**<br>**n(%)** | Māori | 6 (9) | 4 (8) |  | 10 (8) |
|  | NZ European | 22 (32) | 24 (49) |  | 46 (39) |
|  | Pacific Peoples | 20 (29) | 3 (6) |  | 23 (20) |
|  | Indian | 2 (3) | 4 (8) |  | 6 (5) |
|  | Chinese | 4 (6) | 3 (6) |  | 7 (6) |
|  | SE Asian | 2 (3) | 2 (4) |  | 4 (3) |
|  | European /Other | 6 (9) | 9 (18) |  | 15 (12) |
|  | Prefer not to say | 9 (13) |  |  | 9 (7) |
| **PEDSQOL_GIS** |  | mean (SD) | mean (SD) |  |  |
| Total composite score <u>all</u> domains |  | 80.68 (±6.96) | 89.85 (± 4.16) | < .01 | 86.11 (±14.4) |
| *Individual Domain Scores for upper gastroduodenal symptoms only* |  |  |  |  |  |
| Stomach Pain and Hurt (6 items) |  | 73.9 (±23.99) | 83.5 (±17.4) |  | 77.9 (±22.1) |
| Stomach Discomfort When Eating (5 items) |  | 79.8 (±19.40) | 88.3 (±16.2) |  | 83.3 (±18.6)) |
| Food and Drink Limits (6 items) |  | 73.4 (±27.31) | 88.7 (±17.1) |  | 79.7 (±42.8) |
| Trouble Swallowing (3 items), |  | 80.9 (±28.24) | 94.7 (±13.1) |  | 86.6 (±24.2) |
| Heartburn and Reflux (4 items) |  | 76.7 (±20.90) | 87.6 (±18.1) |  | 81.4 (±20.5) |
| Nausea and Vomiting (4 items) |  | 80.5 (±24.51) | 90.9 (±13.2) |  | 84.9 (±21.2) |
| Gas and Bloating (7 items) |  | 75.3 (±21.10) | 87.3 (±21.2) |  | 80.3 (±19.5) |
| Worry about stomach aches (2 items) |  | 79.0 (±29.68) | 90.3 (±17.4) |  | 83.1 (±26.1) |
| Taking Medicines (4 items) |  | 92.8 (±13.22) | 92.9 (±18.9) |  | 91.6 (±18.7) |
| Communication with parents and health professionals (5 items) |  | 94.4 (±16.19) | 97.4 (±7.6) |  | 91.7 (±17.6) |

Overall pictogram translucency and comprehension of the Pediatric and Animation pictogram sets was significantly better than the Adult set in round 1 (78% vs 78% vs 61%, *p* = .03). Comprehension of all pictogram sets increased between rounds 1 and 2, with the most significant increase in the Adult set (61 to 92%, p<.001). Pediatric and Animation sets also improved (78% to 96%, p<0.05). These results are visually represented in the radar plot in Figure 7, showing the three pictogram sets’ comprehension improvement from rounds 1 to 2.

**Figure 7.**
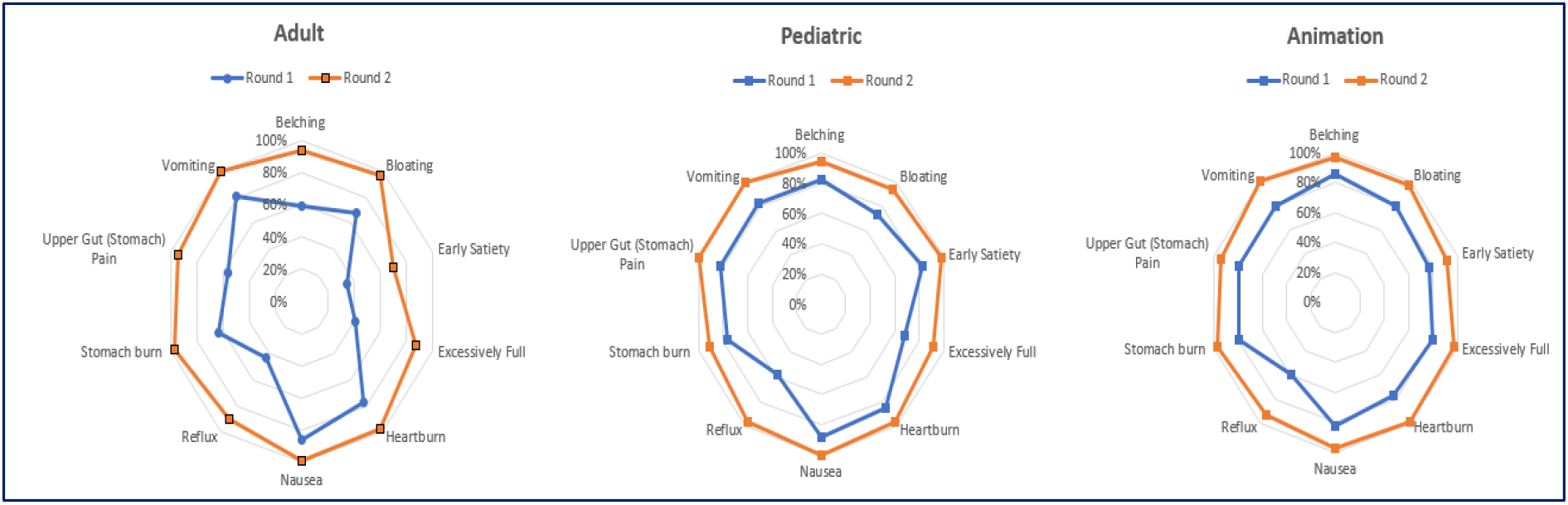
Radar Plot of Comprehension between Rounds 1 and 2 by Symptom and Pictogram Set.

#### The relationship between age and pictogram comprehension

A chi-square test of independence was performed to evaluate the relationship between age and pictogram comprehension in round 1. There was a relationship between age and all pictograms (Adult χ^2^_96_ <.001; Pediatric χ^2^_96_ < .001 and Animation χ^2^_84_ < .001). However, after categorizing age into two age groups (5-12 years and 13-18 years), the relationship was not significant for Adult (χ^2^ _8_ =.29). The relationship remained significant for Pediatric (χ^2^_8_ = .013) and Animation (χ^2^_7_ =.003). The direction of significance, therefore, appeared to favour higher comprehension in the 13-18-year-old participants compared to their younger counterparts.

A further dichotomization of age was undertaken to explore the impact of the very young (5-7 years) on pictogram comprehension. Age data were dichotomized into groups 5-7 years (n=7) and 8-18 years (n=62), and the chi-square test of independence was repeated. There were no changes in the outcomes; however, there was a slight difference in the number of 5-7-year-olds comprehending 5 or more Animation symptoms pictograms (n=5) compared to Pediatric (n=3) and Adult (n=3) pictograms.

#### The relationship between Gastrointestinal Symptoms and Pictogram comprehension

There was no relationship found between the Adult pictogram comprehension and PedsQL-GIS score outcome (Adult χ^2^_8_ =.45). However, there was a significant relationship for Pediatric (χ^2^_8_ < .001) and Animation (χ^2^_7_ < .001) with superior comprehension more likely for participants with more problematic symptoms (lower PedsQL-GIS score).

### Phase 3: Pictogram Preference, Quality and Improvement

There were 118 participants contributing to pictogram preference (General n=69 and BSGM n=49; see Table 1 for combined demographics). Preference for the Pediatric set was significant (χ^2^_2_ < .001). Pictogram preference was consistently higher for each individual Pediatric symptom compared to Adult and Animation (Table 3).

**Table 3.**
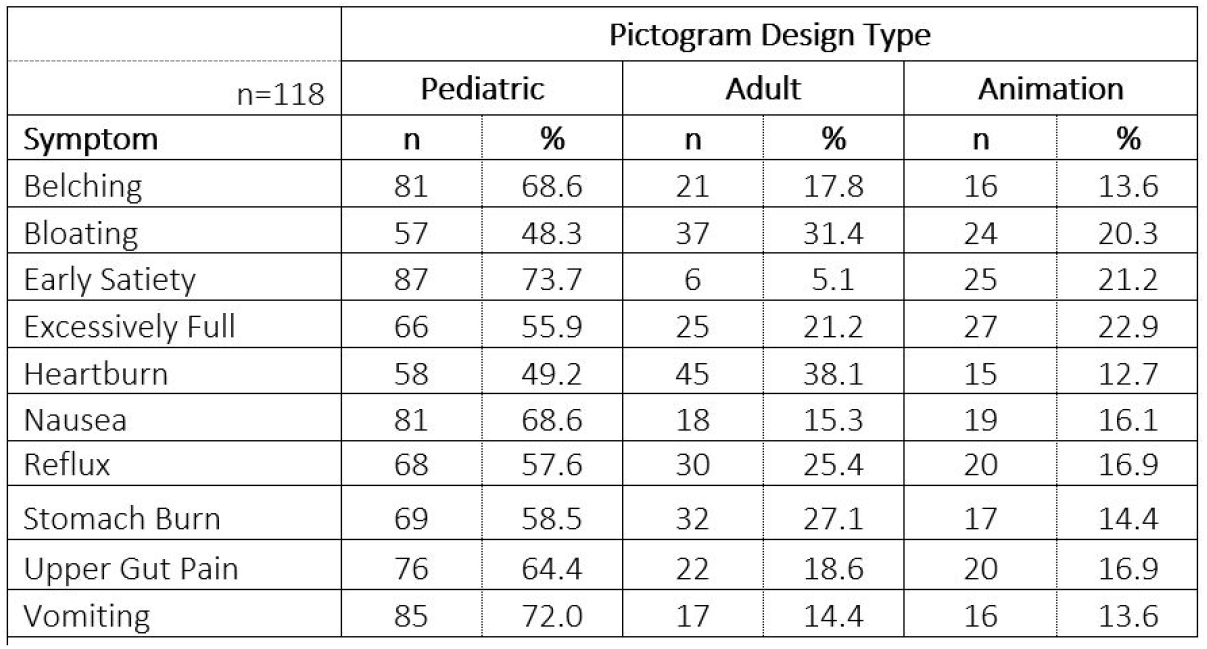
Individual symptom preference by Pictogram design type.

Age was associated with individual symptom preference for Pediatric symptoms of early satiety (χ^2^_28_ = <.001), excessively full (χ^2^_28_ <.001), bloating (χ^2^_28_ = .002), stomach burn (χ^2^_28_ <.001), and heartburn (χ^2^_28_ = .004). Preference for Adult, Pediatric or Animation individual symptoms of upper gut pain, belching, nausea, vomiting and reflux were not significantly related to age.

#### Improvements to the Pictograms

Finally, only twelve participants suggested improving the Pediatric pictograms (all have been adopted). These were confined to emphasizing individual symptom symbolics such as;

“…*increase the size of the flames in heartburn and stomach burn*”,

“…*make the yellowy green bit in the mouth* [for reflux] *brighter as it’s hard to see*,”

“…*need to show more food in the full early one*”.

## DISCUSSION

This paper presents the co-design and validation of a new pediatric set of gastroduodenal symptom pictograms designed for children and young people. There was greater comprehension of and preference for the newly developed Pediatric static pictograms compared to the pre-existing Adult pictograms. Overall, transparency of the pictograms and comprehension in round one for all symptoms was over 80% for the Pediatric and Animation sets, except for excessively full and reflux. These symptom constructs were reported as the least understood in the earlier Adult pictogram validation study (8). Nevertheless, according to the ISO standard testing comprehensibility of graphical symbols (ISO 9186 1 and 2), comprehension by more than 66% of participants is considered validated, increasing to 85% when the subject relates to safety. (11, 12, 23)

The high level of pediatric pictogram comprehension and perceptual quality without text labels or descriptors also supports language independence. This could explain why the Pediatric and Animation pictograms performed well in round 1. This outcome is important as it suggests that these new Pediatric pictograms could reliably capture information from children who may also have literacy difficulties or where English is not their first language. Adding a label and brief symptom description significantly improved comprehension of all pictograms irrespective of age, especially those that were shown to be difficult concepts to comprehend (early satiety, excessively full, reflux).

Little significant additional comprehension was gained by having the Animated pictograms. Comprehension of symptoms using Animation did appear slightly better for children aged 5-7 years, but participant numbers in this age group were small. This finding is supported by previous research that reports that animation can improve comprehension in younger children. (24) There was an association between worse symptomatology and superior comprehension of the Pediatric pictograms. This held for individual symptom pictograms and the Pediatric set overall. Further demonstrating the robustness of the Pediatric pictogram set is that pictogram preference was superior for Pediatric over both Animation and Adult, irrespective of age and comprehension. This suggests that the Pediatric pictograms are relatable and have high perceptual and translucency qualities.

While there are multiple validated questionnaires available for children or their parents to report gastrointestinal symptoms (e.g. Rome IV (25), Peds-Qol-GIS (26), and PROMIS-GI (27)), the complexity of some domain constructs in these questionnaires may lead to difficulty in completing questionnaires with consequent inaccuracies or missed information. (28) Proxy completion by parent or family is an alternative; however, research highlights issues of parent-child alignment. (4, 5) Pictograms are easy to administer digitally or on paper and may have the advantage of greater sensitivity for distinguishing between symptoms. Due to quick application (compared to written questionnaires), pictograms may be more suited to dynamic reporting and may be more able to be used concurrently during physiological investigations or treatments to improve diagnostic information or evaluate treatment efficacy. Previous literature has identified pictograms as a superior way of communicating with patients and patients than text-based content alone. (29, 30)

Despite the similar prevalence of gastroduodenal DGBIs in pediatrics and adults, there is no complete set of validated gastroduodenal symptom pictograms designed with, and for use by, children and young people. (31, 32). Designing and creating any tool, product or service with the target group is a primer for maximizing the chance that the end outcome will function as intended (33, 34). Recognition, understanding and relatability are essential to ensure that the child or young person can identify with the image (35) and the image’s meaning (36). When an image is difficult to interpret, children (and adults) rely on real-world image associations to make sense of the image. They prioritize interpretation through image appearance (37) and draw on real-world experience cues (38). Conceivably, the enhanced stylized elements of the new Pediatric pictograms, including abstract lines to indicate context, emotion, and movement (e.g. the head turned and the hand out in front of a plate of food to depict Early Satiety), helped comprehension. This connection of the pictogram to an association with a real-world referent has been highlighted as an important aspect when supporting understanding, particularly in young children (39). The co-creation of the novel pictograms with participants of different ages and sex into the design of the pictograms has likely contributed to the high relatability, translucency and comprehension outcome. Other research findings report that comprehension of complex constructs is aided when the images provide a sense of self-identification (40) and cultural characteristics. (41)

The static pictograms have the potential to be used digitally or in print, allowing for use in a wide range of settings and for large scale use. The lack of a meaningful difference between the younger and older participants further supports using a single standardized set of pictograms for all children until further research is undertaken in younger children (5-7 years). Therefore, these pictograms will also likely remain relevant for adult use cases, given that the concepts are generalizable symptom expressions that outperformed existing adult reference sets in this study. This approach will limit the potential of introducing a variation of clinical and research outcomes across age groups.

Symptoms of DGBIs often overlap and can wax and wane over me, so diagnosing these disorders is difficult and often frustrating for patients, families, and the clinical team. Without objective criteria nor biomarkers for the underlying causes, symptom assessment plays a significant role in clinical decision-making. Specifically designed, standardized, and validated symptom assessment would enhance the accuracy of symptom reporting and aid in clinical decision-making. Additionally, a clinically applicable tool that is quick and patient-friendly may be applied as a repeated measure or continuous assessment alongside diagnostic investigations or treatments to give more granular information regarding the phenotype of DGBI or the efficacy of the treatment. This novel designed and validated gastroduodenal symptom pictogram set may provide this in clinical and research practice. (42, 43)

The co-design process involved only a small group of young people with relatively homogenous demographics, and the validation studies were performed in a cohort of children who were essentially symptoms naïve. Further validation studies should be considered in pediatric patients with greater symptom burden, a larger younger population, diverse cultural and ethnic groups, and populations from different jurisdictions. Validating the pictograms with more participants with experience of gastroduodenal symptoms will enable concurrent and convergent validity studies to be undertaken.

## Data Availability

Data sharing and data use are governed by the Aotearoa | New Zealand Health and Disability Ethics Committee. The majority of the data is available in the manuscript; however, all requests for additional data can be made to the corresponding author. Requests will be granted if the proposed use aligns with the ethical approval for the study and has obtained relevant ethical approvals, including approval from a NZ Ethical Committee.

## Authorship

GH designed the research study, developed the pictograms, collected and analyzed the data, and wrote the manuscript. CK contributed to the study design, study advice and manuscript revisions. AG contributed to the study design and the final manuscript. CA, HM, and GO’G provided study advice and contributed to the manuscript. All authors peer-reviewed and approved the final version of the manuscript.

